# IMU-derived kinematics detect gait differences with age or knee osteoarthritis but differ from marker-derived inverse kinematics

**DOI:** 10.1101/2022.01.10.22269024

**Authors:** Jocelyn F. Hafer, Julien A. Mihy, Andrew Hunt, Ronald F. Zernicke, Russell T. Johnson

## Abstract

Common in-lab, marker-based gait analyses may not represent daily, real-world gait. Real-world gait analyses may be feasible using inertial measurement units (IMUs), especially with recent advancements in open-source methods (e.g., OpenSense). Before using OpenSense to study real-world gait, we must determine whether these methods: (1) estimate joint kinematics similarly to traditional marker-based motion capture (MoCap) and (2) differentiate groups with clinically different gait mechanics. Healthy young and older adults and older adults with knee osteoarthritis completed this study. We captured MoCap and IMU data during overground walking at participants’ self-selected and faster speeds. MoCap and IMU kinematics were computed with appropriate OpenSim workflows. We tested whether sagittal kinematics differed between MoCap- and IMU-derived data, whether tools detected between-group differences similarly, and whether kinematics differed between tools by speed. MoCap data showed more flexion than IMU data (hip: 0-47 and 65-100% stride, knee: 0-38 and 58-91% stride, ankle: 18-100% stride). Group kinematics differed at the hip (young extension > knee osteoarthritis at 30-47% stride) and ankle (young plantar flexion > older healthy at 62-65% stride). Group-by-tool interactions occurred at the hip (61-63% stride). Significant tool-by-speed interactions were found, with hip and knee flexion increasing more for MoCap than IMU data with speed (hip: 12-15% stride, knee: 60-63% stride). While MoCap- and IMU-derived kinematics differed, our results suggested that the tools similarly detected clinically meaningful differences in gait. Results of the current study suggest that IMU-derived kinematics with OpenSense may enable the valid and reliable evaluation of gait in real-world, unobserved settings.

## 1. Introduction

Gait analyses are a common research tool in biomechanics to detect changes or differences in gait kinematics, which are used as markers of pathology, rehabilitation status, or function (Rodda et al. 2004; Boyer et al. 2012; Curran et al. 2018). While typical laboratory-based gait analyses provide precise, controlled kinematic measurements, they may not accurately represent how people move in uncontrolled, free-living (i.e., real-world) environments. In real-world settings, individuals often walk slower and with shorter stride lengths compared to laboratory settings (Foucher et al. 2010; Hutchinson et al. 2019; Takayanagi et al. 2019; Kim et al. 2020). A mismatch between laboratory-measured gait and real-world gait could lead to incomplete or even spurious conclusions about disease status, intervention success, or function. Thus, accurately and reliably evaluating gait kinematics in real-world, daily life settings is critical.

While traditional, marker-based optical motion capture (MoCap) is challenging to perform in real-world settings (i.e., outside of a lab, without researcher supervision), collecting gait data in free-living environments may be feasible using wearable devices such as inertial measurement units (IMUs). IMUs can be worn by participants as they engage in real-world, daily activities, and many have battery and data storage capabilities that allow for days of continuous data collection. However, the use of IMUs for gait analyses outside the lab is challenging, and therefore few studies exist that assess gait in free-living settings. The challenge of real-world IMU gait analysis is partially because the processing needed to transform IMU data (e.g., sensor-frame angular velocity, linear acceleration, and magnetic heading) into familiar kinematic outcomes (e.g., joint angles) requires data processing methods that are substantially different from those of MoCap gait analyses [e.g., (Seel et al. 2014; Vitali et al. 2017; Dorschky et al. 2019)]. Consequently, many studies use commercially available automated software packages that estimate spatiotemporal outcomes and joint kinematics from IMU data [e.g., (Monda et al. 2015; Nüesch et al. 2017; Washabaugh et al. 2017; Al-Amri et al. 2018)]. While these systems are relatively easy to use, the algorithms are proprietary, and thus a study using one system cannot be replicated or compared against a study using a different system. Additionally, requirements for specific sensor placement and calibration poses of many proprietary systems preclude their use outside of structured, researcher-observed data collection.

Recently, developers at OpenSim released an open-source, freely available software package (OpenSense) to estimate kinematics from IMU data (Seth et al. 2018; Al Borno et al. 2022). OpenSense combines sensor orientation estimates from IMU data with global optimization methods [i.e., inverse kinematics (Lu and O’Connor 1999)] and musculoskeletal models with anatomical constraints to solve for traditional kinematic data like joint and segment angles. This approach may provide the IMU community an accessible, standardized way to report gait kinematics for data collected in non-traditional environments. In contrast to proprietary IMU analysis programs, the parameters and procedures used to process IMU data in OpenSense are published and openly available, therefore the effect of any model assumption on kinematic results can be independently evaluated. Recent studies using OpenSense showed reasonable agreement between IMU and MoCap data for young adult participants (Bailey et al. 2021; Slade et al. 2021; Al Borno et al. 2022).

However, before OpenSense can be used to estimate gait kinematics for clinical populations in real-world settings, we must first demonstrate its ability to detect valid and clinically meaningful (e.g., ≥5°) differences similarly to standard marker-based optical motion capture. Early proof-of-concept studies of OpenSense suggest validity and sensitivity, but only tested healthy young adults (Bailey et al. 2021; Al Borno et al. 2022). Whether this workflow accurately detects true gait differences between populations (e.g., the aged or those with joint pathology) is unknown. Additionally, OpenSense has only been demonstrated in experimental settings where calibration procedures are tightly controlled. Because IMUs do not inherently provide data in reference to anatomy, calibration entails the nontrivial task of transforming data from arbitrary to meaningful reference frames. Existing OpenSense demonstrations either used MoCap measurements in IMU calibration procedures (Al Borno et al. 2022) or adjusted for IMU vs. MoCap model offsets in kinematics results (Bailey et al. 2021). One demonstration did not correct IMU kinematics against MoCap data, but only collected data on a treadmill for young adults (Slade et al. 2021). In order to determine whether OpenSense can give clinically useable estimates of gait kinematics in unobserved, real-world settings, we must evaluate OpenSense kinematics in populations beyond young adults without using calibration procedures that rely on MoCap data.

The aim of this study was threefold. The <u>first aim</u> was to compare IMU- and MoCap-based gait kinematics as calculated using inverse kinematics with the OpenSense toolbox in OpenSim. In keeping with the goal of using OpenSense to estimate kinematics from unobserved, real-world IMU data, we applied IMU calibration procedures without adjusting for differences between IMU model poses and MoCap model poses. Because this approach will ultimately be applied to monitor clinically relevant differences or changes in gait kinematics, we also sought to determine whether OpenSense identified clinically relevant differences across cohorts similarly to optical motion capture methods. Thus, the <u>second aim</u> was to determine whether kinematics calculated from IMU-based and MoCap-based kinematics differed between young adults, older adults, and older adults with knee osteoarthritis (i.e., whether there was a tool-by-group interaction). Finally, because IMU kinematics may be more sensitive to increased speed than MoCap data (Potter et al. 2019), the <u>third aim</u> was to compare kinematics between preferred and faster-than-preferred walking speeds by testing for a tool-by-speed interaction.

## 2. Methods

The current study was a secondary analysis from a previous study (Hafer et al. 2020). These data were used as a convenience sample during a time when human subjects data collection was not feasible (COVID-19 pandemic). Our previous analyses evaluated the validity and reliability of IMU kinematics derived using custom Matlab algorithms.

Those custom algorithms, however, may not be accessible to or easily reproduced by other gait biomechanists, and so in the current study we tested the validity of an open-source, reproducible IMU workflow. Due to the statistical analyses in the current work that required equal numbers of participants per group, we excluded one participant each from the initial study in the young and asymptomatic older adult cohorts.

### 2.1 Protocol summary

Participants included 9 young adults (28.7±4.2 yr), 9 asymptomatic older adults (72.1±3.4 yr), and 9 older adults with knee osteoarthritis (69.2±4.6 yr). Each group included 4 females and 5 males. Participants had BMI <30 kg/m^2^; were able to walk for 30 min without assistive devices; and had no history of major traumatic injury, surgery, or chronic pain in the back or lower extremities (except the knee for the osteoarthritis cohort). Participants with knee osteoarthritis had at least one knee of Kellgren-Lawrence grade II-IV (Kellgren and Lawrence 1957); no surgery within the previous year; and no changes to treatment or joint injections in the previous three months. All participants completed IRB-approved informed consent procedures before completing any study tasks. While the goal of the current work was to demonstrate feasibility for IMU gait analyses outside of lab settings, this proof-of-concept was conducted using overground walking trials collected in a laboratory to be able to compare with simultaneous traditional MoCap data.

We collected gait data from participants’ right leg (for young and asymptomatic older adults) or the leg with the greater radiographic osteoarthritis severity (for older adults with knee osteoarthritis). We recorded 10 trials of MoCap and IMU data simultaneously as participants walked overground at preferred and faster-than-preferred (verbal instruction: “walking to catch the bus” (Brinkerhoff et al. 2019)) speeds. Participant characteristics and basic spatiotemporal measures are shown in Table 1.

**Table 1.** Participant characteristics and walking speeds

|  | Young |  | Older healthy |  | Older with knee osteoarthritis |  |
| --- | --- | --- | --- | --- | --- | --- |
|  | Mean | SD | Mean | SD | Mean | SD |
| n (male/female) | 9 (5/4) |  | 9 (5/4) |  | 9 (5/4) |  |
| Age (years) | 28.7 | 4.2 | 72.1 | 3.4 | 69.2 | 4.6 |
| Height (m) | 1.70 | 0.08 | 1.70 | 0.08 | 1.73 | 0.12 |
| Bodymass (kg) | 71.1 | 8.3 | 70.8 | 9.1 | 74.8 | 12.1 |
| Preferred walking speed (m/s) | 1.36 | 0.17 | 1.33 | 0.15 | 1.18 | 0.08 |
| Faster walking speed (m/s) | 1.83 | 0.31 | 1.85 | 0.19 | 1.78 | 0.23 |

### 2.2 Inertial Measurement Unit Methods

We affixed IMU sensors (Opal v2, APDM Inc., Portland, OR, USA) to each participant’s sacrum, midpoint of the lateral thigh, midpoint of the lateral shank, and dorsal foot. Thigh and shank sensors were placed on participants’ skin with double-sided tape and then secured with elastic wrap.

IMU sensors captured data at 128 Hz. Sensors were synchronized to each other and collected data continuously for the duration of the session. IMU data collection occurred simultaneously with MoCap data capture, but the two systems were not synchronized in real time. We recorded the beginning and end of walking trials using an IMU event trigger (one trial = one walk across the MoCap volume). Using trigger events, we split continuous IMU data into walking trials in MATLAB. We extracted one stride from each walking trial based on gait events identified using a one-dimensional continuous wavelet transform of the foot vertical acceleration data (Baroudi et al. 2020; Hafer et al. 2020). Finally, we used a Kalman filter to estimate sensor orientations in a global reference frame from raw linear acceleration, angular velocity, and magnetometer signals (Holmstrom 2016 Oct 26) and exported the IMU orientation data in quaternion format.

We used the OpenSim “gait2354” model (Delp et al. 2007) to calculate kinematics. The model had virtual IMU sensors placed such that the axes for each sensor approximated the orientation of the native axes of each sensor when a participant stood in a neutral posture. For each walking trial, we created a new calibrated model from a frame of static, neutral posture data that immediately preceded the walking trial. In accordance with our goal to evaluate the potential for this workflow in unobserved setting, the calibration frame was identified from raw sacrum sensor accelerometer and gyroscope signals alone. The OpenSense calibration procedure registers the relative orientation of a sensor’s data in the global reference frame to an assumed model pose, effectively giving a sensor-to-model offset. In the current study, participants were assumed to be in a neutral posture immediately before beginning to walk.

For each trial of IMU data, we used the calibrated model (i.e., one model per trial for IMU data) to calculate the generalized coordinates using the inverse kinematics algorithm in OpenSim (Lu and O’Connor 1999). We used equal tracking weights for all segments and did not use heading correction. Analyses focused on sagittal plane variables, and as such extracted the pelvic tilt segment angle, and hip flexion, knee flexion, and ankle flexion joint angles from the inverse kinematics analyses for each trial. All kinematic data were time-normalized to 0-100% of the gait cycle.

### 2.3. Optical Motion Capture Methods

To compare IMU data to MoCap data, we used a standard marker set to model the pelvis and leg. The marker set included anatomical markers on the right and left anterior and posterior superior iliac spines, right and left greater trochanters, medial and lateral femoral epicondyles and malleoli, first and fifth metatarsal heads, and lateral and medial heel (note—foot markers were placed on shoes). Rigid clusters of four non-collinear markers each were affixed to both the thigh and shank. We collected MoCap data with 15 cameras (Vicon, Oxford UK) capturing at 120 Hz and 2 force plates (AMTI, Watertown MA) capturing at 1200 Hz.

We used Visual3D (c-motion, Germantown, MD USA) to perform initial processing of MoCap data. We first low-pass filtered marker data at 8 Hz and then identified gait events (e.g., heel strike and toe off) automatically using force plate contact. Subsequently, we exported marker and force data for a static calibration trial and any walking trial containing a stride of data with clean force plate contact (at least 4 strides per condition for each participant). After exporting from Visual3D, we used a MATLAB pipeline to rotate the data to match the OpenSim coordinate system convention and convert files to the OpenSim format.

After converting marker data to OpenSim format, we scaled the gait2354 model (Delp et al. 2007) for each participant using participant body mass and the marker data from the static calibration trial. We then modified each scaled model by locking the subtalar joint, such that the only degree-of-freedom at the ankle was flexion. The pelvis, thigh, shank, and foot were scaled based on anatomical markers on these segments (i.e., bilateral ASIS and PSIS markers, greater trochanter, medial and lateral femoral epicondyles, medial and lateral malleoli, 1^st^ and 5^th^ metatarsal heads and calcaneus).

For each trial, we used the subject-scaled model (one model per subject for all marker data) to calculate the generalized coordinates using the inverse kinematics algorithm. We then extracted the same sagittal-plane coordinates (pelvic tilt, hip flexion, knee flexion, and ankle flexion angles) for comparison with the IMU data. All kinematic data were time-normalized to 0-100% of the gait cycle.

### 2.4 Statistical Analysis

Due to different gait event identification methods by tool (i.e., force plate detection for MoCap versus acceleration-based detection for IMU), we expected small offsets in the relative timing of MoCap and IMU data. To remove this processing artifact from our comparisons, we corrected for inter-tool timing differences for each speed condition before running statistical analyses. To do this, we first assumed that the timing of the peak knee flexion angle during the gait cycle should be the same regardless of tool. We identified the difference in the timing of the peak knee flexion angle between MoCap and IMU trials. Then, we time-shifted IMU data forward or backward such that the peak knee flexion angle occurred at the same percentage of the gait cycle for both IMU and MoCap data. Data were then averaged within each tool and speed for each participant, such that statistical analyses included four time series per participant per joint (i.e., MoCap and IMU at preferred and faster speeds). Subject means were used for statistical comparisons, because the statistical method use required the same number of trials for each subject and condition. We compared time-series waveforms for pelvic tilt and hip, knee, and ankle flexion across tools (MoCap vs. IMU), groups (young, healthy older, vs. older with knee osteoarthritis), and walking speeds (preferred vs. faster) with a continuous 3-way ANOVA implemented using statistical parametric mapping [SPM, (Pataky 2010)]. When we found significant group effects or significant interactions, we used t-tests implemented in SPM for post-hoc comparisons.

## 3. Results

MoCap data showed more anterior pelvic tilt than IMU data (0-100% gait cycle, p<0.001) and more flexion than IMU data across joints (hip: 0-38 and 60-100% gait cycle; knee: 0-38 and 57-89% gait cycle; ankle: 6-99% gait cycle; all differences p<0.001) (Figure 1, Table 2). Young adults had greater hip extension than the knee osteoarthritis group during midstance (24-48% gait cycle, p<0.001, Figure 2). Critically, group by tool interactions were minimal (56-65% gait cycle at hip, p=0.024) and did not overlap with main effects of group.

**Table 2.** Statistical results for all comparisons. RMSD in tool comparison indicates mean and range root mean squared difference between MoCap and IMU data for each joint. Percentages (%s) indicate ranges of the gait cycle found to be significantly different via SPM. Post-hocs indicate direction of main effect or interaction effect differences. Not sig. indicates a main, interaction, or post-hoc effect was not significant. n/a indicates a post-hoc comparison was not run because a main or interaction effect was not significant.

|  |  | Tool |  |  |  | Group |  |  |  |  |  | Speed |  |  |  |
| --- | --- | --- | --- | --- | --- | --- | --- | --- | --- | --- | --- | --- | --- | --- | --- |
|  | All variables in ° | IMU |  | Marker |  | Young |  | Older Healthy |  | Knee Osteoarthritis |  | Preferred |  | Faster |  |
|  |  | Mean | SD | Mean | SD | Mean | SD | Mean | SD | Mean | SD | Mean | SD | Mean | SD |
| **Pelvis** | Average tilt | 1.1 | 3.1 | -4.6 | 4.0 | 0.1 | 2.7 | -2.5 | 3.0 | -2.8 | 2.4 | -1.1 | 2.1 | -2.4 | 2.2 |
| **Hip** | Peak extension | 14.8 | 3.3 | 11.9 | 7.7 | 15.0 | 3.0 | 14.6 | 4.6 | 10.1 | 4.0 | 12.2 | 14.6 | 14.0 | 17.0 |
|  | ROM | 41.9 | 6.0 | 44.8 | 7.3 | 42.8 | 4.6 | 44.7 | 5.0 | 39.8 | 6.5 | 39.1 | 3.4 | 44.6 | 4.3 |
| **Knee** | Peak flexion | 54.3 | 7.6 | 69.9 | 4.7 | 62.5 | 4.2 | 63.3 | 4.3 | 60.5 | 6.3 | 61.4 | 21.0 | 62.9 | 20.8 |
|  | ROM | 60.5 | 6.1 | 70.3 | 6.8 | 66.2 | 4.9 | 66.9 | 6.7 | 62.5 | 6.0 | 64.2 | 5.4 | 65.6 | 3.6 |
| **Ankle** | Peak plantar flexion | 25.3 | 6.9 | 12.3 | 7.0 | 23.9 | 4.0 | 16.0 | 4.5 | 15.0 | 7.2 | 17.4 | 7.9 | 20.2 | 8.1 |
|  | ROM | 32.6 | 6.9 | 24.5 | 4.9 | 31.9 | 2.8 | 26.1 | 4.4 | 25.5 | 6.6 | 27.3 | 3.2 | 28.4 | 3.1 |

**Figure 1.**
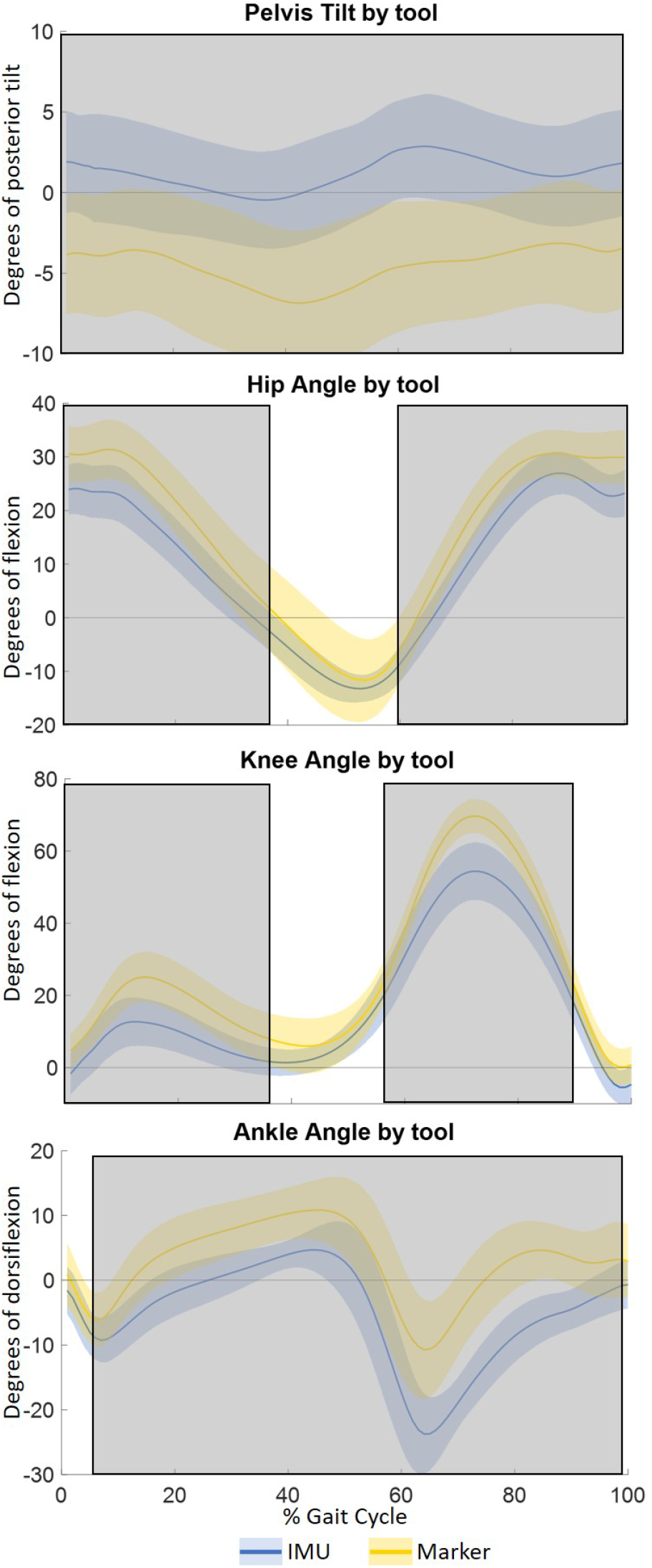
Sagittal pelvis, hip, ankle, and knee angles across tools (mean±SD). Shaded grey rectangles indicate difference between tools (p<0.05) by statistical parametric mapping.

**Figure 2.**
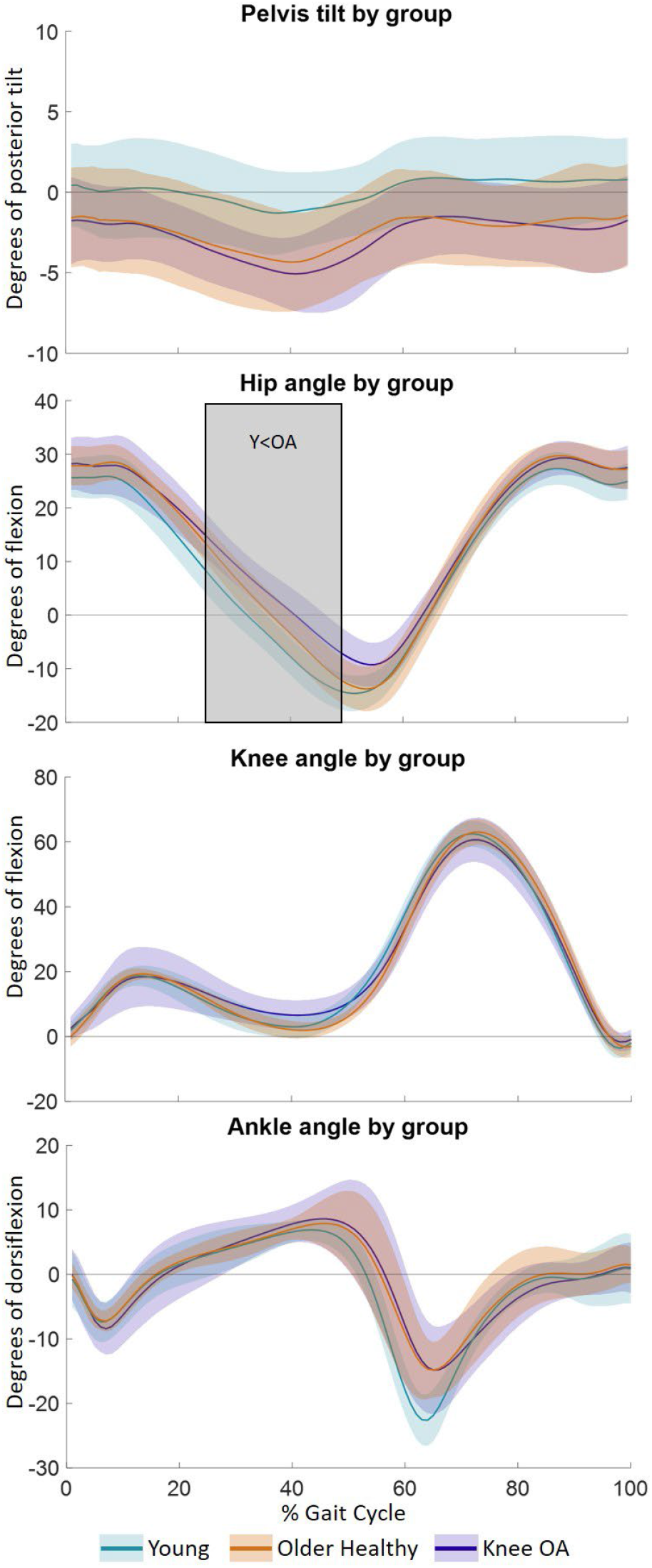
Sagittal pelvis, hip, ankle, and knee angles across groups (mean±SD). Shaded grey rectangles indicate significant main effect of group (p<0.05) by statistical parametric mapping with accompanying text indicating post-hoc group differences.

As expected [e.g., (Fukuchi et al. 2019)], there were significant effects of speed on kinematics (Figure 3, Table 2). At the faster walking speed, the pelvis was more anteriorly tilted (1-94% gait cycle, p<0.001) and the hip was more flexed at 0-22 and 60-100% gait cycle (p<0.01) and more extended at 36-52% gait cycle (p<0.01). At the faster speed, the knee was more flexed from 2-21 and 51-71% gait cycle (p=0.001) and more extended from 81-91% gait cycle (p=0.018). Compared to preferred speed, at the faster speed, ankle dorsiflexion was greater from 1-3, 6-31, and 67-100% gait cycle (p<0.05) and ankle plantar flexion was greater from 43-63% gait cycle (p<0.001). There were significant tool-by-speed interactions at the pelvis and all three joints, with faster speed causing greater increases in hip flexion for marker data than IMU data (12-15% gait cycle, p=0.04). There were no significant group-by-tool-by-speed interactions. Full statistical results can be found in Table 3.

**Table 3.** Representative discrete kinematics by tool, group, and speed. All results reported in degrees and absolute value, except pelvis tilt where positive values indicate posterior tilt and negative values indicate anterior tilt.

|  | Tool | Group |  | Speed | Tool×Group |  | Tool×Speed |  |
| --- | --- | --- | --- | --- | --- | --- | --- | --- |
|  |  | Main | post-hoc |  | Main | post-hoc | Main | post-hoc |
| **Pelvic tilt** | 0-100% (p<0.001) | not sig. | n/a | 3-53% (p<0.001)<br>59-94% (p=0.003) | not sig. | n/a | 0-5% (p=0.048)<br>28-100% (p<0.001) | not sig. |
| **Hip angle** | 0-47% (p<0.001)<br>65-100% (p<0.001) | 30-47% (p=0.005) | Y<OA 30-47%<br>(p<0.001) | 0-22% (p<0.001)<br>37-52% (p=0.01)<br>60-100% (p<0.001) | 61-63% (p=0.046) | not sig. | 12-34% (p=0.002) | MoCap F>P 12-15% (p=0.04)<br>IMU not sig. |
| **Knee angle** | 0-38% (p<0.001)<br>58-91% (p<0.001) | not sig. | n/a | 5-21% (p=0.003)<br>52-71% (p=0.001)<br>81-92% (p=0.014) | not sig. | n/a | 16-18% (p=0.046)<br>60-63% (p=0.045)<br>85-90% (p=0.037) | MoCap F>P 60-63% (p=0.007)<br>and 88-90% (p=0.048)<br>IMU F>N 60-63% (p=0.028) |
| **Ankle angle** | 18-100% (p<0.001) | 62-65% (p=0.039) | Y<OH 61-65%<br>(p=0.023) | 1-30% (p<0.001)<br>42-64% (p<0.001)<br>67-100% (p<0.001) | not sig. | n/a | 11% (p=0.05) | not sig. |

**Figure 3.**
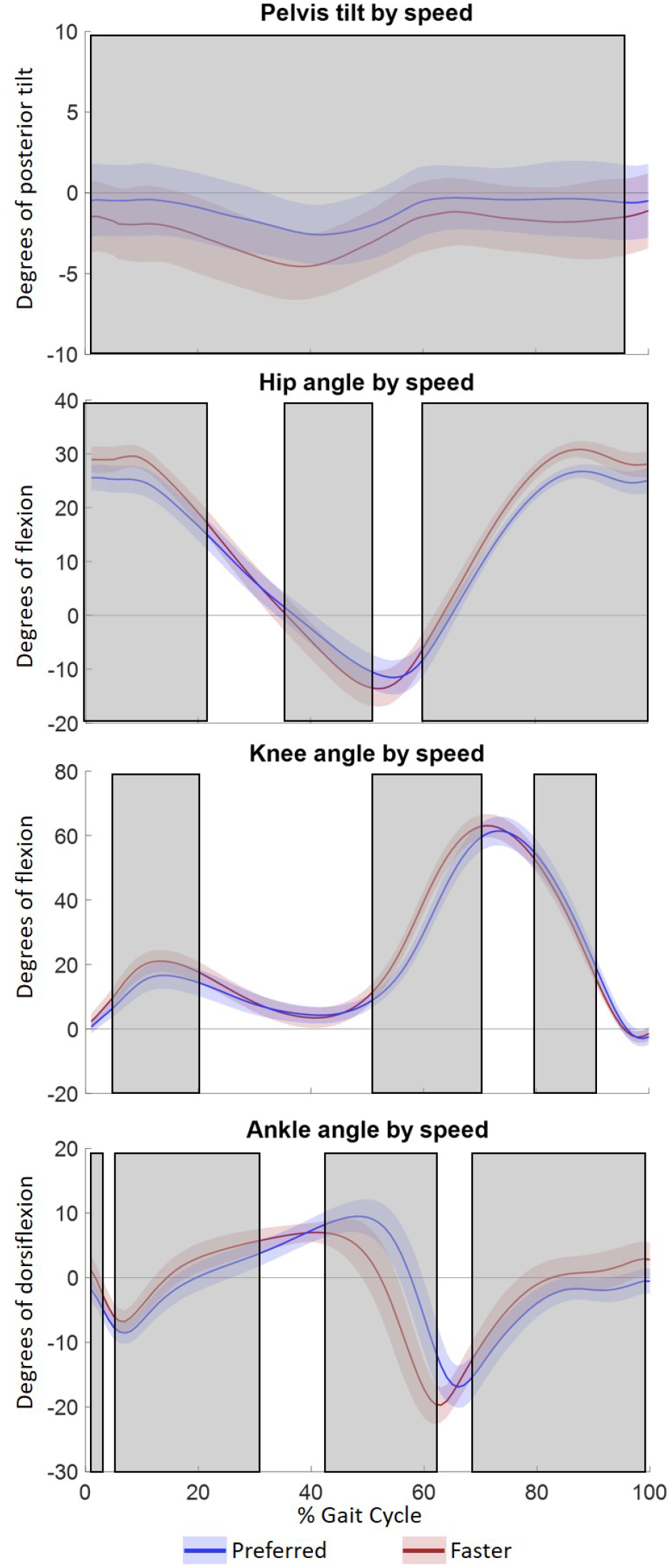
Sagittal pelvis, hip, ankle, and knee angles across speeds (mean±SD). Shaded grey rectangles indicate significant main effect of speed (p<0.05) by statistical parametric mapping.

## 4. Discussion

The ability to reliably measure kinematics outside the lab with IMUs would expand understanding of how age, pathology, or interventions affect real-world gait. Thus, the current study aimed to determine whether lower extremity sagittal kinematics differed when derived from traditional MoCap or IMUs using a workflow that could theoretically be applied outside the lab, and whether differences across young adults, older healthy adults, and older adults with knee osteoarthritis could be detected similarly with both tools. We found that lower extremity kinematics were significantly different between MoCap- and IMU-derived data, but between-group differences were similar for both tools (i.e., minimal group × tool interaction). These results suggest that, while the absolute kinematics derived from these two systems differ, the relative differences in kinematics derived using IMU data with OpenSense may enable detection and tracking of clinically relevant gait differences.

When comparing MoCap and IMU kinematics, we saw apparent offsets between tools (Figure 1) as well as potential differences in the dynamic tracking between tools (i.e., different range of motion between tools in Table 3). Our findings differ from other recent studies that found minimal differences in kinematics between MoCap and IMU data using an OpenSim workflow (Bailey et al. 2021; Slade et al. 2021; Al Borno et al. 2022). We found slightly larger differences between MoCap and IMU kinematics (average joint RMSD 6-9°, Table 2) than previous studies that used OpenSense (RMSD 3-7°) (Bailey et al. 2021; Slade et al. 2021; Al Borno et al. 2022). These between-study differences are likely largely due to different model calibration procedures (i.e., apparent offset between MoCap and IMU data in Figure 1). In the current study, we used periods of static data as calibration time points, assuming that participants were standing in a neutral posture at these times. Any differences in participant posture from neutral or between assumed IMU alignment and actual anatomical reference frames would therefore be carried forward into inverse kinematics results. In contrast, Al Borno et al. and Bailey et al. corrected for differences between MoCap and IMU model postures and Slade et al. carefully controlled participant posture during calibration data capture (Bailey et al. 2021; Slade et al. 2021; Al Borno et al. 2022). Compared to a measured or visually verified IMU calibration posture, the assumed IMU calibration postures would likely introduce some differences between IMU and MoCap-derived kinematics. However, if researchers or clinicians plan to implement an OpenSense workflow in unobserved, real-world data collections, measured or carefully controlled calibration postures would not be available. Thus, the current results provide a more realistic estimate of the validity of OpenSense kinematics in unobserved data collections.

The between-group hip extension differences in the current study align with age- or knee osteoarthritis-related differences in previous studies. Similar to previous findings in older adults and adults with knee osteoarthritis (Astephen et al. 2008; Boyer et al. 2017), in the current study, older adults with knee osteoarthritis had ∼5° less peak hip extension and 2° less hip range of motion compared to young adults (Table 3). Qualitatively, discrete knee and ankle kinematics suggest that the knee osteoarthritis group had less knee range of motion compared to the healthy groups (62.7±6.5° for OA vs. 66.8±4.3° and 67.0±6.7°) and that both older groups had less peak ankle plantar flexion compared to the young group (16.2±5.1° and 15.6±6.8° vs. 23.1±3.9°; Table 3). The agreement between the group differences in the current study and in previous studies—despite absolute differences between IMU and MoCap kinematics—support the use of IMU-derived data for detecting or tracking clinically meaningful differences in sagittal plane gait kinematics.

Unsurprisingly, sagittal plane kinematics differed between preferred- and faster-than-preferred walking speeds. The current study, however, was focused on whether speed affected kinematics differently for MoCap and IMU data. Integrated IMU data (e.g., linear velocity and displacement and angular orientation) are susceptible to greater error when noise increases at greater movement speed. While this error is generally low at walking speeds (Potter et al. 2019), OpenSense does not directly implement error-reducing procedures such as linear drift correction or stride-by-stride integration resetting. We did find significant tool × speed interactions at every joint (Table 2), but hose interactions appeared to have little clinical relevance as post-hoc comparisons of the effect of speed on MoCap or IMU data revealed that MoCap resulted in greater hip flexion than IMU for only 4% of the gait cycle at faster speeds. Thus, IMU data appeared—overall—to detect similar speed-related differences in kinematics as MoCap data overall.

To date, many validations of IMU kinematic data have relied on proprietary commercial software packages, which have RMSD of 3-10° between MoCap and IMU sagittal kinematics during controlled, in-lab data collection (Nüesch et al. 2017; Kobsar et al. 2020). While proprietary software may provide internally reliable and accurate estimates of kinematics, the analysis methods within those software packages are unknown, making comparison across studies or replication of results with different systems difficult or impossible. Our results suggest that the open-source OpenSense analysis tool may enable kinematic measurement with similar accuracy to proprietary software, even without strict calibration procedure. This type of open-source software package provides an opportunity to standardize IMU kinematic processing across different studies and increases accessibility of IMU analyses to researchers and clinicians who do not have the expertise or resources to develop customized data processing algorithms.

The current study has a couple potential limitations that could be expanded upon in future work. Most importantly, demonstration of the validity of this IMU approach in a lab setting does not guarantee similar validity in real-world, unobserved settings. While we attempted to mimic some aspects of unobserved data collection (e.g., selecting calibration time points based on IMU signal characteristics rather than known, controlled postures; identifying gait events from IMU signals), we were able to control for other unobserved data challenges, including detection of walking activity or sensor placement variation. While many methods exist for activity classification and gait event detection [e.g., (Attal et al. 2015; Benson et al. 2019; Gurchiek et al. 2020)], the impact of variations in sensor placement remains a relatively understudied area. As part of the development of this study, we examined the effect of variation in sensor placement on IMU kinematic outcomes. We found that differences in sensor placement (i.e., ∼2-4 cm superior/inferior position or ∼90° medial rotated position) could impart differences in peak sagittal joint angles of up to 10° (Supplemental material). This finding may support the use of additional IMU data alignment techniques prior to inputting data to OpenSense to minimize the effect of day-to-day or inter-participant sensor placement differences.

The assumptions and constraints of the chosen musculoskeletal model in the current study may have impacted the ability to detect some expected group differences. In our previous analysis of these data (Hafer et al. 2020), we found significant differences in knee range of motion between the young and knee osteoarthritis cohorts that were not seen in the current work. For simplicity in the current study, we used a model with one degree of freedom knee joint rotation (Yamaguchi and Zajac 1989). In combination with a sagittal-only ankle joint, this model may have compensated for true frontal or transverse plane static offsets or gait patterns in unexpected ways. In our previous work, where we used a functional alignment method to calibrate IMU data with no assumptions of linked segments, we found no significant differences in knee range of motion calculated using MoCap or IMU data (Hafer et al. 2020). In the current study, knee and ankle range of motion estimates differed between MoCap and IMU kinematics by 10 and 7°, respectively. Examining the sensitivity of kinematics to modelling choices and static vs. functional IMU data alignment may help elucidate ways to minimize these inter-tool differences.

Overall, this study demonstrates that an open-source, freely available IMU inverse kinematics approach (OpenSense) can provide estimates of gait kinematics that can differentiate between clinically relevant groups in controlled collection settings.

This work is a foundational step in determining whether an inverse kinematics approach sufficiently detects or tracks meaningful decrements or improvements in real-world gait function. Real-world demonstration of these methods will be essential to verify that these in-lab findings translate to unobserved data collections.

## Data Availability

All data produced in the present study are available upon reasonable request to the authors. Some data and code used in this work are available at https://simtk.org/projects/knee-oa-age-imu.

## Data Availability

Sample data and code are available at https://simtk.org/projects/knee-oa-age-imu

## Supplemental Material

A supplemental file demonstrating an analysis of the effect of differences in IMU sensor placement on inverse kinematics results is included with this article.

