## Supplemental file for "IMU-derived kinematics detect gait differences with age or knee osteoarthritis but differ from marker-derived inverse kinematics"

**Effects of IMU sensor placement on OpenSense inverse kinematics**

<u>Overview and Brief Summary</u>

We tested whether kinematics of the pelvis, hip, knee, and ankle differed if sensors were placed in different locations on lower limb segments. We tested locations that might be commonly recommended sensor placements (e.g., dorsal foot vs. heel) or that might represent likely variations in sensor locations if non-experts placed sensors themselves (e.g., superior-inferior sensor location difference or anterior-posterior sensor location difference on thigh or shank).

We found that there were minor to moderate differences in joint kinematics across sensor placements (Figures 3-5). The differences were beyond what we would expect from inter-sensor differences in noise characteristics alone and were most apparent around kinematic peaks.

<u>Sensor placement comparison procedure</u>

We collected overground gait data using a pelvis sensor placed over the sacrum and three sensors per segment on the right thigh, shank, and foot (Figure 1).

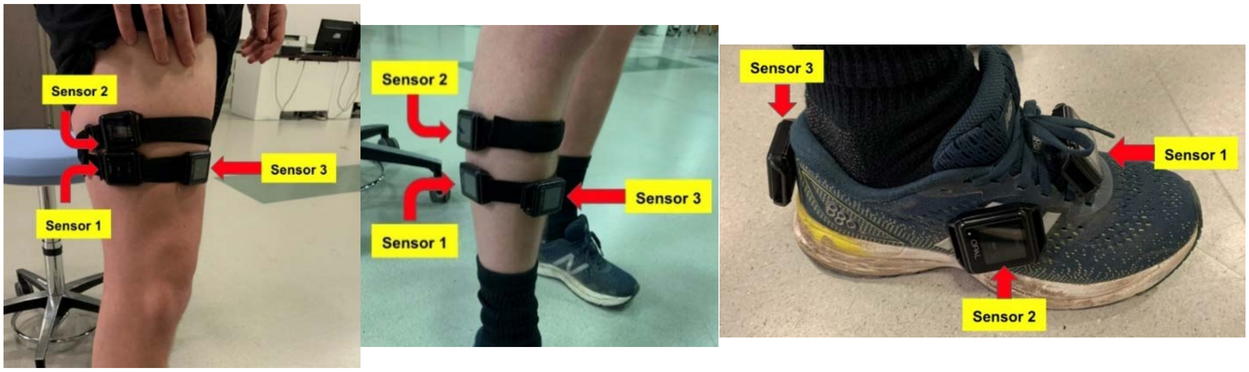


**Figure 1.** Placements for thigh, shank, and foot sensors

We used the IMU Placer tool and the same frame of static data to create 7 separate calibrated models incorporating each sensor configuration. Sensor axis orientations varied as expected in each calibrated model (Figure 2).

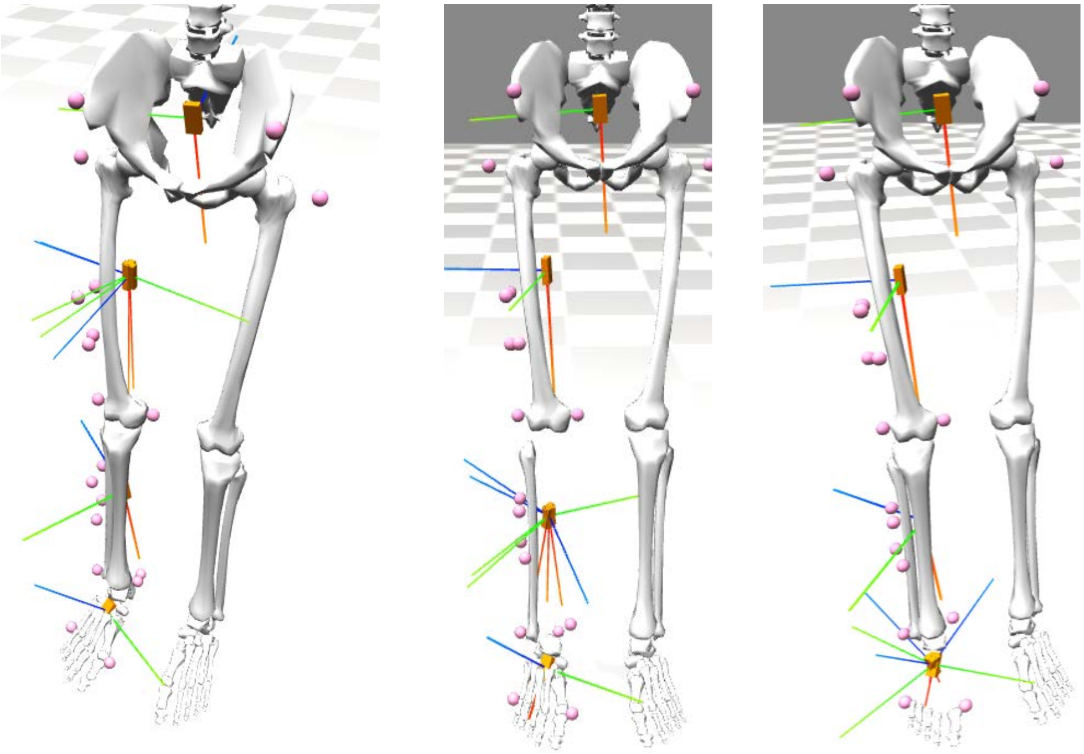

**Figure 2.** Calibrated model sensor axis orientations for standard sensor placements overlaid with 2 other thigh sensor placements (left), 2 other shank sensor placements (middle), and 2 other foot sensor placements (right)

We next ran inverse kinematics for each of the calibrated models using the same range of walking data (with appropriate sensor data included for each model). Kinematics showed expected ranges and patterns (accounting for a not-standardized static calibration pose), with some variation between sensor placements. For results shown below, IK was run with a weighting of 1 for all sensors. Each figure below shows the difference in inverse kinematics results between placements for a single sensor (e.g., the “Thigh” figure shows the effect of different thigh sensor placements). Blue lines show results for all standard sensor placements (all sensors sensor 1 from Figure 1). Orange lines show results when the thigh, shank, or foot sensor was in position 2. Yellow lines show results when the thigh, shank, or foot sensor was in position 3. Note that the same pelvis sensor placement was used for all comparisons, and so pelvis data do not vary.

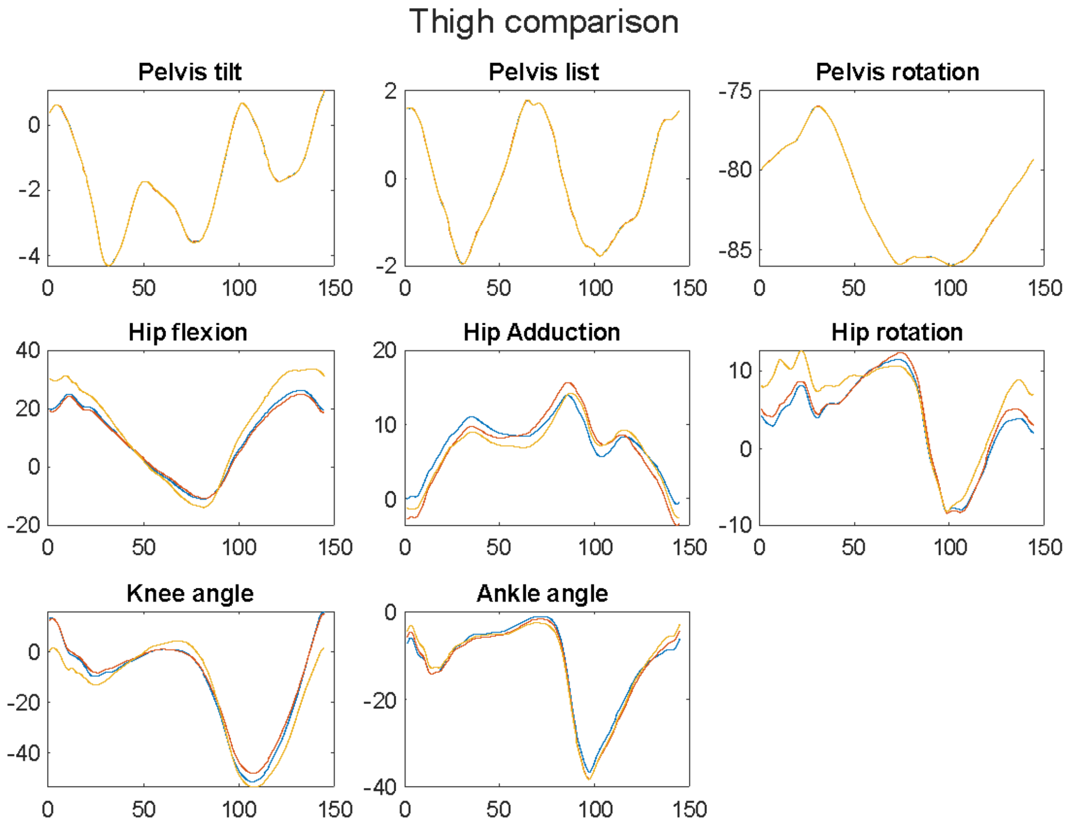


**Figure 3.** Effect of different thigh sensor placements on inverse kinematics results. Blue = Thigh sensor 1; Orange = Thigh sensor 2; Yellow = Thigh sensor 3. All other sensors always in position 1.

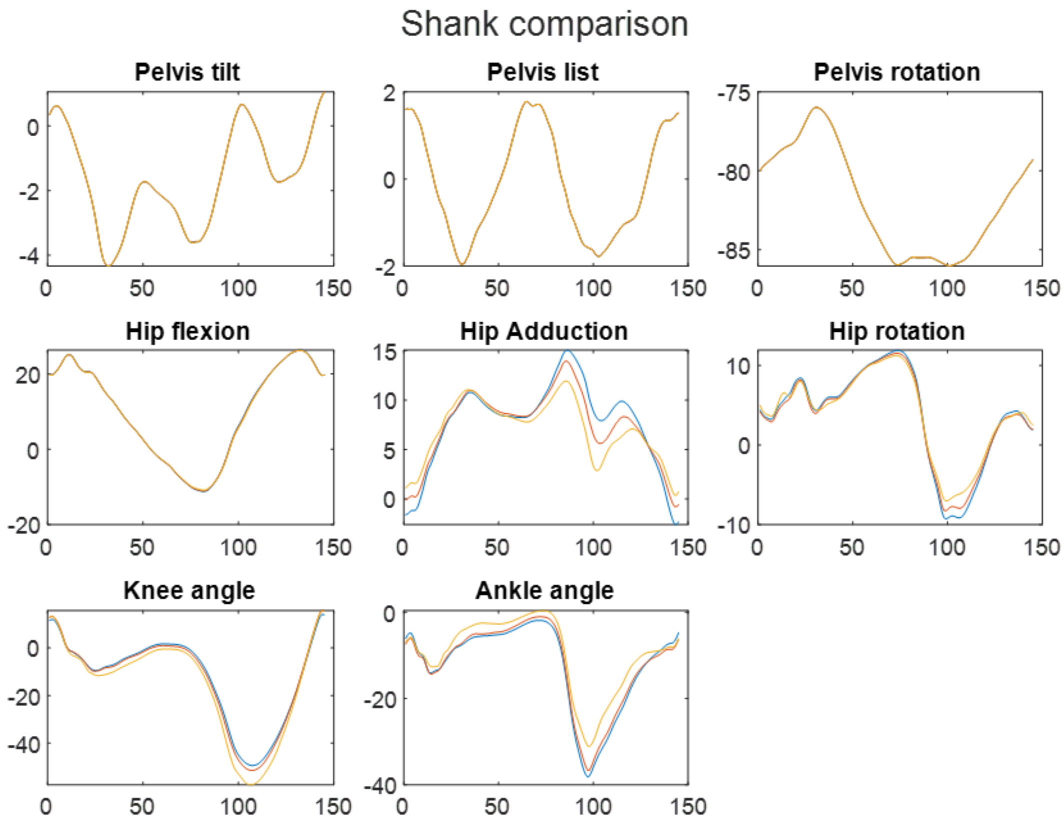


**Figure 4.** Effect of different shank sensor placements on inverse kinematics results. Blue = Shank sensor 1; Orange = Shank sensor 2; Yellow = Shank sensor 3. All other sensors always in position 1.

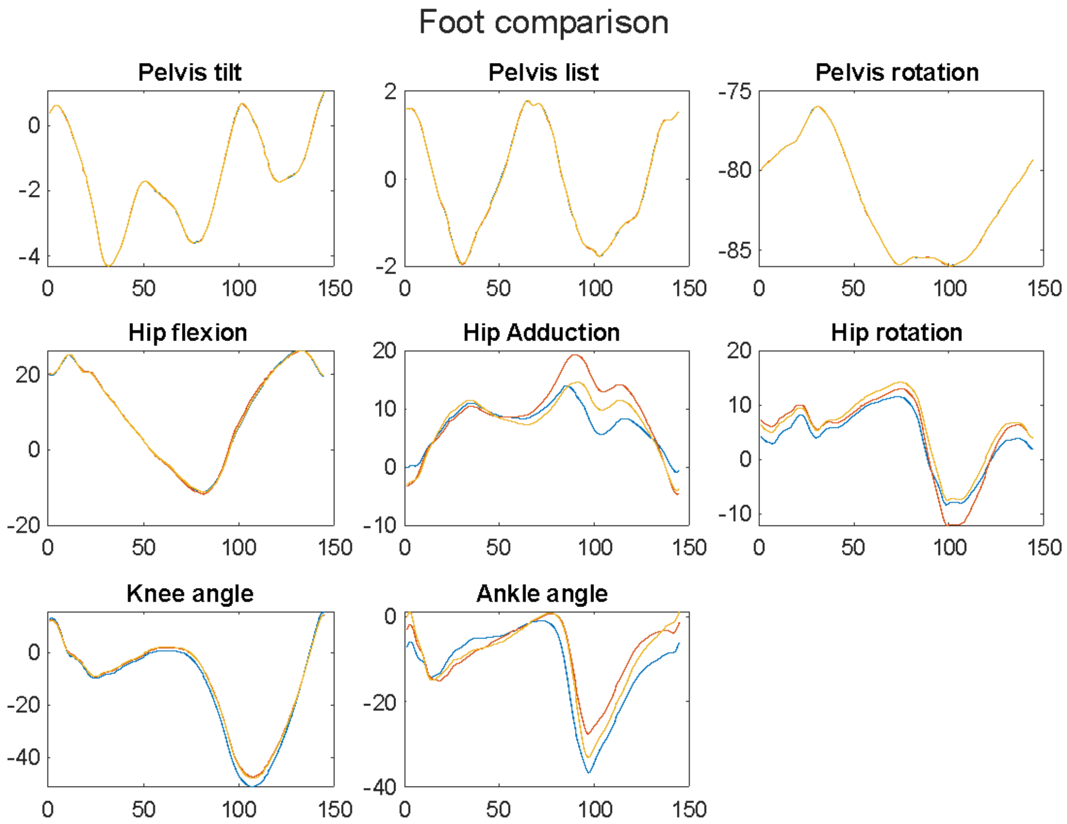


**Figure 5.** Effect of different foot sensor placements on inverse kinematics results. Blue = Foot sensor 1; Orange = Foot sensor 2; Yellow = Foot sensor 3. All other sensors always in position 1.
